# COVID-19 OUTCOMES IN MS: EARLY EXPERIENCE FROM NYU MULTIPLE SCLEROSIS COMPREHENSIVE CARE CENTER

**DOI:** 10.1101/2020.05.12.20094508

**Authors:** Erica Parrotta, Ilya Kister, Leigh Charvet, Carrie Sammarco, Valerie Saha, R.E. Charlson, Jonathan Howard, Josef Maxwell Gutman, Malcolm Gottesman, Nada Abou-Fayssal, Robyn Wolintz, Marshall Keilson, Cristina Fernandez-Carbonell, Lauren B. Krupp, Lana Zhovtis Ryerson

## Abstract

**Objective:** Report outcomes on patients with Multiple Sclerosis (MS) and related disorders with COVID-19 illness.

**Methods:** From March 16 to April 30^th^, 2020, patients with MS or related disorders at NYU Langone MS Comprehensive Care Center (MSCC) were identified with laboratory-confirmed or suspected COVID-19. The diagnosis was established using a standardized questionnaire or by review of in-patient hospital records.

**Results:** We identified 76 patients (55 with relapsing MS of which 9 had pediatric-onset;17 with progressive MS; and 4 with related disorders). 37 underwent PCR testing and were confirmed positive. Of the entire group, 64 (84%) patients were on disease-modifying therapy (DMT) including anti-CD20 therapies (n=34, 44.7%) and sphingosine 1-phosphate receptor modulators (n=10, 13.5%). The most common COVID-19 symptoms were fever and cough, but 21.1% of patients had neurologic symptom recrudescence preceding or coinciding with the infection. A total of 18 (23.7%) were hospitalized; 8 (10.5%) had COVID-19 critical illness or related death. Features more common among those hospitalized or with critical illness or death were older age, presence of comorbidities, progressive disease, and a non-ambulatory status. No DMT class was associated with an increased risk of hospitalization or fatal outcome.

**Conclusions:** Most MS patients with COVID-19 do not require hospitalization despite being on DMTs. Factors associated with critical illness were similar to the general ‘at risk’ patient population. DMT use did not emerge as a predictor of poor COVID-19 outcome in this preliminary sample.

## Background

At present, we do not know if multiple sclerosis (MS) or disease-modifying therapies (DMT) for MS increase the risk of acquiring COVID-19 or worsen the course (hospitalizations, ICU, death). DMT medications have immunosuppressive effects that could hamper mounting an effective immune response to the infection.^1^ On the other hand, immunosuppression could offer protection by downregulating hyperinflammation and the cytokine storm associated with severe disease.^2^

New York City emerged as the epicenter of the COVID-19 pandemic in the United States in March 2020. Given the widespread prevalence of COVID-19 in our community, clinicians at the NYU Multiple Sclerosis Comprehensive Care Center (MSCCC) received numerous reports of COVID-19 infection from patients and began systematically collecting symptom data and the clinical course. This timely, real-world observational study on outcomes of COVID-19 in actively treated MS patients could inform clinicians as they counsel MS patients and guide treatment decisions during the pandemic.

## Methods

Demographic and clinical features were collected on patients currently followed at MSCCC and its 4 affiliated sites in the greater New York area (2 in Long Island, 2 in Brooklyn) with a history of COVID-19 infection from March 16th through April 30, 2020. All patients who contacted the center with infectious symptoms or were seen during routine teleneurology visits were queried regarding COVID-19 exposure using a standardized instrument. Inclusion criteria were any patient with MS or related disorders who was diagnosed with COVID-19 by a healthcare provider (based on symptoms, course, radiographic findings consistent with CDC COVID-19 criteria^3^ and/or positive COVID-19 PCR when available). For hospitalized patients, in-patient records were reviewed. NYU School of Medicine Institutional Review Board approval was obtained for the study.

Descriptive statistics were used to summarize the demographic and clinical characteristics of patients. Categorical variables were summarized as counts and percentages. No imputation was made for missing data. Differences between hospitalized patients were compared by t-test, χ2 test, or Fisher’s exact test where appropriate.

## Results

A total of 76 patients met inclusion criteria, 72 (93%) with MS, and 4 (7%) with related disorders. Average age was 44.9±15.2 years (range 13-71) and 61.8% were female. Disease duration was 15.2±10.7 years (range 1-52). Racial breakdown was: 50 (65.8%) White, 21 (27.6%) Black, 3 (3.9%) Asian, 1 (1.3%) Pacific Islander, and 1 (1.3%) other. Hispanic ethnicity was reported by 15 (19.7%). Of the 72 MS patients, 55 (76.4%) had a relapsing-remitting subtype and 17 (23.6%) primary- or secondary-progressive subtypes. Diagnoses of four non-MS patients were: neuromyelitis optica spectrum disorder (NMOSD, n=1), chronic-relapsing-inflammatory-optic-neuropathy (CRION, n=1), neurosarcoidosis (n=1), and myelin-oligodendrocyte-glycoprotein-IgG associated disorder (MOGAD, n=1). 65 patients (84%) were on DMT.

Common symptoms reported included fever (68.4%), cough (68.4%), fatigue (38.2%), shortness of breath (31.6%), and myalgias/arthralgias (26.3%). Other frequent symptoms included anosmia (22%), ageusia (19.7%), and headache (21.1%). A subset reported neurologic symptom recrudescence (21.1%) suggestive of relapse. In some cases, neurologic symptoms preceded viral symptoms by several days.

Of the 84% of patients on DMTs, 44.7% were treated with anti-CD20 therapies (rituximab n =18; ocrelizumab n= 16), 13.5% on Sphingosine-1-phosphate (S1P) modulators (fingolimod n= 8; siponimod n=2), 7.9% (n=6) on glatiramer acetate, 5.3% (n=4) each on natalizumab and dimethyl fumarate and 3.9% (n=3) on beta-interferons. As shown in Table 1, there were no significant differences between DMT use among those who were and were not hospitalized (p=0.75) or between those specifically treated with anti-CD20 therapies (43% vs 50%, p=0.40). Table 1 summarizes outcomes by DMT class.

**Table 1:** DISEASE MODIFYING THERAPIES SPECIFIC OUTCOMES.

| **Demographics** | Anti-CD20 | S1P inhibitors | Glatiramer acetate | Natalizumab | Dimethyl fumarate | Interferon | IVIG | None |
| --- | --- | --- | --- | --- | --- | --- | --- | --- |
| n | 34 | 10 | 6 | 4 | 4 | 3 | 3 | 12 |
| Age, years (SD) | 38.72 (SD 14.9) | 44.90 (SD 11.2) | 53.3 (SD 14.8) | 37.8 (SD 17.9) | 52.8 (SD 7.2) | 57.6 (SD 16.7) | 57 (SD 12.8) | 49.8 (SD 15.8) |
| Female n (%) | 20 (62.5%) | 6 (60%) | 4 (66.7%) | 2 (50%) | 2 (50%) | 1 (33.3%) | 3 (100%) | 8 (66.7%) |
| **Ambulation status, n (%)** |  |  |  |  |  |  |  |  |
| Ambulatory | 27 (79.4%) | 9 (90%) | 2 (33.3%) | 4 (100%) | 3 (75%) | 3 (100%) | - | 8 (66.7%) |
| Ambulatory with assistance | 4 (11.8%) | 1 (10%) | 3 (50%) | - | - | - | 2 (66.7%) | 1 (8.3%) |
| Non-ambulatory | 3 (8.8%) | - | 1 (16.7%) | - | 1 (25%) | - | 1 (33.3%) | 3 (25%) |
| **Hospitalization** |  |  |  |  |  |  |  |  |
| Not hospitalized | 25 (73.5%) | 9 (90%) | 5 (83.3%) | 3 (75%) | 2 (50%) | 3 (100%) | 3 (100%) | 8 (66.7%) |
| Hospitalized | 9 (26.5%) | 1 (10%) | 1 (16.7%) | 1 (25%) | 2 (50%) | - | - | 4 (33.3%) |
| **Comorbidities associated with COVID-19** |  |  |  |  |  |  |  |  |
| Obesity | 9 (26.5%) | 3 (30%) | 2 (33.3%) | 2 (50%) | 1 (25%) | - | 1 (33.3%) | 6 (50%) |
| Hypertension | 4 (11.8%) | 3 (30%) | 1 (16.7%) | 1 (25%) | 2 (50%) | - | 2 (66.7%) | 4 (33.3%) |
| Diabetes | 1 (2.9%) | 1 (10%) | 1 (16.7%) | 1 (25%) | 1 (25%) | - | 1 (33.3%) | 2 (16.7%) |
| CAD | 1 (2.9%) | - | 1 (16.7%) | 1 (25%) | - | - | - | - |
| **Covid-19 Outcomes** |  |  |  |  |  |  |  |  |
| Recovered | 24 (70.6%) | 7 (70%) | 4 (66.7%) | 3 (75%) | 4 (100%) | 3 (100%) | 2 (66.7%) | 8 (66.7%) |
| Death | 2 (5.8%) | - | 1 (16.7%) | 1 (25%) | - | - | - | 2 (16.7%) |

Legend: Anti-CD20: ocrelizumab, rituximab; S1P inhibitors: fingolimod, siponimod; Interferons: interferon beta-1a, interferon beta-1b; IVIG intravenous immunoglobulins. One 58-year-old female with history of hypertension on leflunomide had a mild course.

As shown in Table 2, of the full sample, 18 (23.7%) were hospitalized. The hospitalized vs. non-hospitalized patients were more likely to be older (51.9 vs 42.7 years), have progressive MS subtype (38.8% vs 17.2%), be less ambulatory (50% vs. 19% requiring assistance or non-ambulatory, and have comorbid obesity (61.1% vs 20.6%, p=0.002), coronary artery disease (16.7% vs 0%, p=0.01) and/or history of venous thromboembolism (16.7% vs 1.7%, p=0.04).

**Table 2:** DEMOGRAPHICS OF HOSPITALIZED VS NON-HOSPITALIZED COVID-19 PATIENTS.

| ***Demographics*** | **Hospitalized**<br>**(n=18)** | **Non-Hospitalized**<br>**(n=58)** | **p value\*** |
| --- | --- | --- | --- |
| Age, years (range, SD) | 52.0 (17-71, 14.6) | 42.7 (13-71, 14.8) | .028 |
| Sex, (% Female) | 10 (55.6%) | 37 (63.8%) | 0.36 |
| Race, n (% white) | 12 (66.7%) | 38 (65.5%) | 0.35 |
| Ethnicity, n (% Hispanic) | 6 (33.3%) | 9 (15.5%) | 0.20 |
| ***Clinical diagnosis*** |  |  |  |
| RRMS | 8 (44.4%) | 47 (81.0%) | .004* |
| Progressive MS | 7 (38.8%) | 10 (17.2%) | 0.06 |
| Other Demyelinating Syndrome | 3 (16.7%) | 1 (1.7%) | 0.03 |
| Disease duration, years<br>(range, SD) | 17.2 (5- 36, 10.5) | 14.6 (1-52, 10.8) | 0.40 |
| ***Ambulation Status, n (%)*** |  |  |  |
| Ambulatory | 9 (50%) | 47 (81%) | 0.01 |
| Ambulatory with assistance | 5 (27.8%) | 6 (10.3%) | 0.08 |
| Non-ambulatory | 4 (22.2%) | 5 (8.6%) | 0.12 |
| ***Comorbidities associated with COVID-19*** |  |  |  |
| Obesity | 11 (61%)(%) | 12 (20.7%) | 0.003* |
| Hypertension | 5 (27.8%) | 12 (20.6%) | 0.37 |
| Coronary artery disease | 3 (16.6%) | - | 0.01 |
| Diabetes | 2 (11.1%) | 6 (10.3%) | 0.61 |
| Venous Thromboembolism | 3 (16.6%) | 1 (1.7%) | 0.04 |
| ***COVID-19 Outcomes*** |  |  |  |
| Death, n (%) | 5 (27.7%) | 1 (1.7%) | 0.002 |
| Recovered, n (%) | 9 (50.0%) | 47 (81.0%) | — |

Legend: \*Descriptive comparisons based on independent sample t-test, Pearson Chi Square or Fisher's Exact; significance set at .004 to correct for multiple comparisons

Eight patients (10.5%) had critical illness defined by ICU admission (n=5) and/or death (n=6). Compared to the entire MS sample, the critically ill patients were older (mean of 57.7±10.5, range 42-71), more likely to have a progressive subtype (50.0%), and less ambulatory (62.5% requiring assistance or non-ambulatory). Following the pattern observed in those who were hospitalized, the critically ill group had had high rates of comorbid obesity (62.5%), coronary artery disease (25%), and venous thromboembolism (37.5%). A detailed table of these patients has been compiled and may be available upon request, and/or pending peer review journal acceptance.

Nine patients with pediatric-onset MS were identified as the center has a large pediatric MS sample (Table 3). Ages ranged from 13 to 26 years; 8 were female and all had relapsing-remitting MS. Noted comorbidities were obesity (n=3), Type I diabetes (n=1), or both (n=1). Two of the nine were hospitalized requiring supplemental oxygen. None required invasive ventilation. Eight patients were either fully recovered or recovering at censoring date. One remains hospitalized.

**Table 3:**
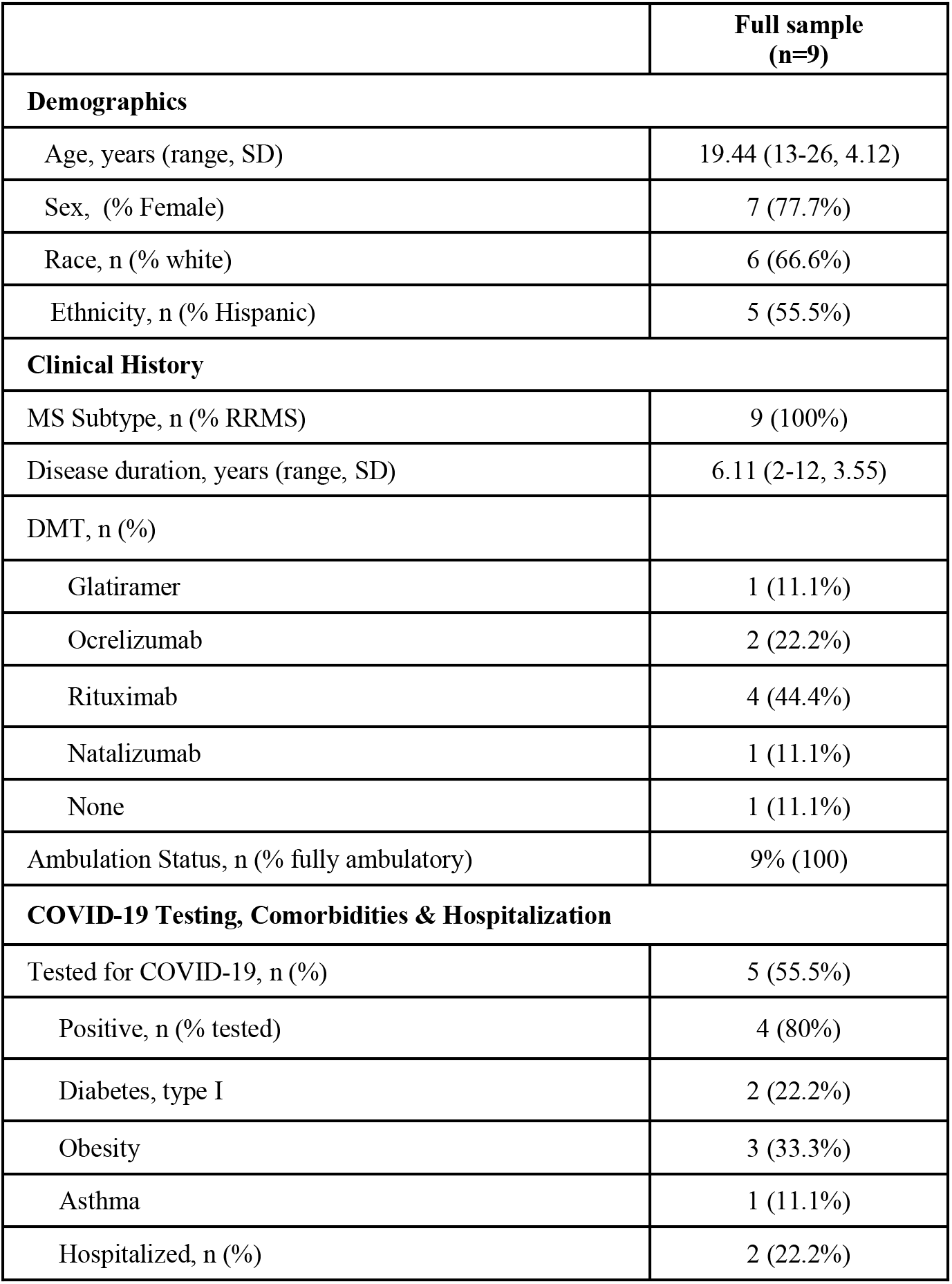
DEMOGRAPHICS OF PEDIATRIC ONSET MULTIPLE SCLEROSIS PATIENTS WITH COVID-19.

|  | Full sample<br>(n=9) |
| --- | --- |
| **Demographics** |  |
| Age, years (range, SD) | 19.44 (13-26, 4.12) |
| Sex, (% Female) | 7 (77.7%) |
| Race, n (% white) | 6 (66.6%) |
| Ethnicity, n (% Hispanic) | 5 (55.5%) |
| **Clinical History** |  |
| MS Subtype, n (% RRMS) | 9 (100%) |
| Disease duration, years (range, SD) | 6.11 (2-12, 3.55) |
| DMT, n (%) |  |
| Glatiramer | 1 (11.1%) |
| Ocrelizumab | 2 (22.2%) |
| Rituximab | 4 (44.4%) |
| Natalizumab | 1 (11.1%) |
| None | 1 (11.1%) |
| Ambulation Status, n (% fully ambulatory) | 9% (100) |
| **COVID-19 Testing, Comorbidities & Hospitalization** |  |
| Tested for COVID-19, n (%) | 5 (55.5%) |
| Positive, n (% tested) | 4 (80%) |
| Diabetes, type I | 2 (22.2%) |
| Obesity | 3 (33.3%) |
| Asthma | 1 (11.1%) |
| Hospitalized, n (%) | 2 (22.2%) |

## Discussion

Patients with MS and related disorders often seek guidance regarding the impact of the disease and medication on their risk of COVID-19 infection. This observational study provides some insights regarding COVID-19 disease course and outcomes in actively treated patients with MS and related disorders.

The rate of hospitalization in our patients (24%) and mortality (7.9%) are in-line with the data of other published MS patient database^4^ and for the general population of patients with COVID-19.^5^ Similar risk factors were identified and included older age, male sex, and higher number of comorbidities identified including obesity, diabetes, hypertension, and coronary artery disease^6-8^. MS-specific features associated with more severe COVID-19 included non-ambulatory status and progressive disease course. Given the small sample size, we could not determine whether these patients were at higher risk given their advanced age and other comorbidities, or whether worse disability in and of itself represents an additional risk factor for complications.

We did not observe an association between DMT class and COVID-19 outcome in univariate comparisons, however, our sample size is small and these preliminary findings should be interpreted with caution. There is a relatively high proportion of COVID-19 infected patients on anti-CD20 therapies (44.7%) compared to our MSCCC population in which 33.1% of patients take anti-CD20 therapies. This observation can be an artifact of data collection (such patients are more likely to have regular visits during this period) or might represent an increased susceptibility to COVID-19 infection, as has been noted with psoriasis patients on other biologic therapies^9^ and non-COVID-19 infections in MS patients on anti-CD20 therapies in general.^1^

As a note of caution in the care of MS patients, a subgroup of individuals reported worsening of preexisting neurologic symptoms before or at onset of COVID-19 symptoms. It is important to consider COVID-19 in the differential of worsening neurological symptoms and to be cautious about the indiscriminate use of steroids.

Our study has several limitations. This was a convenience sample and not randomly selected, nor was the entire practice systematically surveyed. Patients were identified during routine teleneurology visits, if they notified the office, or if hospitalized. This likely led to an over-representation of more symptomatic individuals. Although we used a systematic questionnaire to collect relevant data, we could not verify this data independently unless the patient was seen by NYU Langone-affiliated physician or hospital. Compounding this problem was lack of access to COVID-19 testing in our area. Less than half of our patients (48.7%) underwent SARS-COV2 PCR testing. As shown in Table 4, subgroup analysis showed that this group was not different from the overall sample with respect to demographic or MS-related features.

**Table 4:** DEMOGRAPHICS & OUTCOMES OF ALL PATIENTS VS COVID-19 PCR CONFIRMED POSITIVE.

|  | All patients (n=76) | COVID PCR Positive (n=37) |
| --- | --- | --- |
| **Demographics** |  |  |
| Age, years (range, SD) | 44.9 (13-71, 15.2) | 47.5 (13-71, 15.15) |
| Sex, F/M (% Female) | 47/29 (61.8%) | 23/14 (62.2%) |
| **Race, n (%)** |  |  |
| White | 50 (65.8%) | 24 (64.9%) |
| Black | 21 (27.6%) | 12 (32.4%) |
| Asian | 3 (3.9%) | - |
| Pacific Islander | 1 (1.3%) | - |
| Other | 1 (1.3%) | 1 (2.7%) |
| **Ethnicity, n (%)** |  |  |
| Not-Hispanic | 59 (77.6%) | 28 (75.7%) |
| Hispanic | 15 (19.7%) | 8 (21.6%) |
| Other | 2 (2.6%) | 1 (2.7%) |
| **Smoking Status, n (%)** |  |  |
| Never | 52 (68.4%) | 25 (67.5%) |
| Former | 20 (26.3%) | 10 (27.02) |
| Current | 2 (2.6%) | 1 (2.7%) |
| Unknown | 2 (2.6%) | 1 (2.7%) |
| **Clinical** |  |  |
| **Multiple Sclerosis, n (%)** |  |  |
| RRMS | 55 (72.3%) | 25 (67.6%) |
| SPMS | 15 (19.7%) | 10 (27.0%) |
| PPMS | 2 (2.6%) | - |
| Other Demyelinating Syndrome, n (%) | 4 (5.2%) | 2 (5.4%) |
| Disease duration, years (range, SD) | 15.2 (1-52, 10.7) | 16.9 (2-52, 11.46) |
| **DMT, n (%)** |  |  |
| Anti-CD20 | 34 (44.7%) | 14 (37.8%) |
| S1P inhibitors | 10 (13.2%) | 4 (10.8%) |
| Glatiramer acetate | 6 (7.9%) | 4 (10.8%) |
| Natalizumab | 4 (5.3%) | 3 (8.1%) |
| Dimethyl fumarate | 4 (5.3%) | 2 (5.4%) |
| Leflunomide | 1 (1.3%) | 1 (2.7%) |
| Interferons | 3 (3.9%) | 1 (2.7%) |
| IVIG | 3 (3.9%) | 1 (2.7%) |
| None | 12 (15.8%) | 7 (18.9%) |
| **Ambulation Status, n (%)** |  |  |
| Ambulatory | 56 (73.7%) | 24 (64.9%) |
| Ambulation with assistance | 11 (14.4%) | 7 (18.9%) |
| Non-ambulatory | 9 (11.8%) | 6 (16.2%) |
| **Comorbidities associated with COVID-19** |  |  |
| Hypertension | 17 (22.4%) | 11 (29.7%) |
| Obesity | 23 (30.3%) | 11 (29.7%) |
| Diabetes | 8 (10.5%) | 5 (13.5%) |
| Venous thromboembolism (VTE) | 4 (5.3%) | 3 (8.1%) |
| Coronary artery disease (CAD) | 3 (3.9%) | 3 (8.1%) |
| History of Cancer | 4 (5.3%) | 2 (5.4%) |
| Baclofen pump | 3 (3.9%) | 2 (5.4%) |
| Indwelling foley | 3 (3.9%) | 2 (5.4%) |
| None | 24 (31.6%) | 12 (32.4%) |
| **COVID-19 Outcomes** |  |  |
| Death, n (%) | 6 (7.9%) | 5 (13.5%) |
| Recovered, n (%) | 56 (73.7%) | 25 (67.6%) |

Legend: Anti-CD20: Ocrelizumab, Rituximab; S1P inhibitors: fingolimod, siponimod; Interferons: interferon beta-1a, interferon beta-1b; IVIG intravenous immunoglobulin

Our early experience with COVID-19 at NYU Langone MSCCC could inform clinicians taking care of MS patients during the pandemic. Our findings suggest that individuals with MS who experience COVID-19 have similar disease course, outcomes, and risk factors for complications as the general population. Rigorous, population-based studies are needed to confirm our preliminary findings and better define the risk of COVID-19 infection with respect to individual DMTs.

## Data Availability

I have full access to all of the data and have the right to publish any and all data separate and apart from any sponsor

